# Optimal Pre-dilatation Treatment before Implantation of a Magmaris Bioresorbable Scaffold in Coronary Artery Stenosis. The OPTIMIS trial

**DOI:** 10.1101/2024.07.26.24311089

**Authors:** Kirstine Nørregaard Hansen, Jens Trøan, Akiko Maehara, Manijeh Noori, Mikkel Hougaard, Julia Ellert-Gregersen, Karsten Tange Veien, Anders Junker, Henrik Steen Hansen, Jens Flensted Lassen, Lisette Okkels Jensen

**Affiliations:** Department of Cardiology, Odense University Hospital, Odense, Denmark; University of Southern Denmark, Odense, Denmark; Cardiovascular Research Foundation, New York Presbyterian Hospital, New York, USA

## Abstract

**Introduction:** Bioresorbable scaffolds (BRS) have been developed to overcome limitations related to late stent failures of drug-eluting-stents, but previous studies have observed lumen reduction over time after implantation of BRS. The aim of the study was to investigate if lesion preparation with a scoring balloon compared to a standard non-compliant balloon minimizes lumen reduction after implantation of a Magmaris BRS (MgBRS) assessed with optical coherence tomography (OCT) and intravascular ultrasound (IVUS).

**Method:** Eighty-two patients with stable angina pectoris were included and randomized in a ratio 1:1 to lesion preparation with either a scoring balloon or a standard non-compliant balloon prior to implantation of a MgBRS. The primary endpoint was minimal lumen area (MLA) 6 months after MgBRS implantation.

**Results:** Following MgBRS implantation, MLA (6.4 ± 1.6 mm^2^ vs. 6.3 ± 1.5 mm^2^, p=0.65), mean scaffold area (7.8 ± 1.5 mm^2^ vs. 7.5 ± 1.7 mm^2^, p=0.37), and mean lumen area (8.0 ± 1.6 mm^2^ vs. 7.7 ± 2.1 mm^2^, p=0.41) did not differ significantly in patients where the lesions were prepared with scoring vs. standard non-compliant balloon respectively. Six-month angiographic follow-up with OCT and IVUS was available in seventy-four patients. The primary endpoint, 6-months MLA, was significantly larger in lesions prepared with a scoring balloon compared to a standard non-compliant balloon (4.7 ± 1.4 mm^2^ vs. 3.9 ± 1.9 mm^2^, p=0.04), whereas mean lumen area (7.2 ± 1.4 mm^3^ vs. 6.8 ± 2.2, p=0.35) did not differ significantly. IVUS findings showed no difference in mean vessel area at the lesion site from baseline to follow-up in the scoring balloon group (16.8 ± 2.9 mm^2^ vs. 17.0 ± 3.6 mm^2^, p=0.62), whereas mean vessel area (17.1 ± 4.4 mm^2^ vs. 15.7 ± 4.9 mm^2^, p<0.001) was smaller in lesions prepared with a standard non-compliant balloon due to negative remodeling.

**Conclusion:** Lesion preparation with a scoring balloon prior to implantation of a MgBRS resulted in significantly larger MLA after 6 months due to less negative remodeling compared to lesion preparation with a standard non-compliant balloon.

Registration: URL: https://www.clinicaltrials.gov; Unique identifier: NCT04666584.

**Clinical perspectives:** What is new?

- Intense lesion preparation with a scoring balloon prior to implantation of a magnesium-based Magmaris bioresorbable scaffold results in less lumen reduction and malapposition after 6 month compared to conventional lesion preparation with a non-compliant balloon in patients with stable angina.
- Negative remodeling was seen in lesions treated with conventional lesion preparation, whereas optimal lesion preparation with a scoring balloon caused in stable remodeling.

What are the Clinical Implications?

- Lesions preparation with a scoring balloon is safe and ensures better vascular healing and vessel dynamics after implantation of a magnesium-based Magmaris bioresorbable scaffold.
- Optimal lesion preparation should be considered before implantation of magnesium-based Magmaris bioresorbable scaffold.

## Introduction

Bioresorbable scaffolds (BRS) were developed to provide temporary vessel support during the early phases of coronary vessel healing, leaving the artery stent-free after degradation as an alternative to drug-eluting stents (DES) during percutaneous coronary intervention (PCI)^1, 2^. The potential advantages of BRS were restored vasomotion and potential reduction in late stent failures. The Absorb everolimus-eluting BRS (Abbott Vascular, Abbott Park, IL, USA) showed increased risk of scaffold thrombosis and vessel shrinkage over time^3^ with significant minimal lumen area (MLA) reduction after 6 months assessed with optical coherence tomography (OCT)^4^. It is hypothesized that the mechanism behind lumen reduction is based on decreased radial strength in BRS compared with bare-metal stents and risk of recoil and scaffold dismantling^5^. The construction of BRS continued to develop, and different types are now available on the market. The magnesium-based BRS (Magmaris, Biotronik, Bülach, Switzerland) (MgBRS) was later introduced with improved radial strength, stronger backbone, change in drug-polymer coating and showed better efficacy compared to the first BRSs^6–9^. Head-to-head comparison between newer generation DES and the MgBRS is limited, but the anti-restenotic efficacy has not yet solved the scaffold failure^5, 10^.

Optimal lesion preparation prior to implantation of a MgBRS appeared to facilitate optimal scaffold sizing and better expansion post-procedure in complex lesions^11^, but the effect of aggressive pre-dilation on vessel and lumen changes over time is uncertain. Peri-procedural intravascular imaging is recommended during implantation of a MgBRS due to lack of a radiolucent backbone. OCT is ideal to assess lumen contours^12^, whereas intravascular ultrasound (IVUS) provides information on the vessel wall and vessel remodeling over time^13, 14^. The aim of this study was to assess whether a more aggressive lesion preparation with a scoring balloon compared to a standard non-compliant balloon prior to implantation of a MgBRS resulted in less lumen reduction MLA after 6 months.

## Methods

### Study design

The OPTIMIS (Optimal pre-dilatation Treatment before Implantation of a Magmaris bioresorbable scaffold In coronary artery Stenosis) study was a prospective, randomized-controlled trial conducted at Odense University Hospital in Denmark from December 2020 to September 2023. The study compared lesion preparation with a scoring balloon to a standard non-compliant balloon, prior to implantation of a MgBRS and the effect on lumen dimension in the scaffold treated segment after 6 months. The patients were randomized to the two pre-dilatation methods in a ratio 1:1. The primary hypothesis of the OPTIMIS-study was that intense lesion preparation with a scoring balloon prior to implantation for a MgBRS would result in a larger MLA after 6-month follow-up, compared to standard pre-dilatation with a non-compliant balloon. A detailed description of the study design has previously been published^15^.

The study was approved by the Regional Committees on Health Research Ethics for Southern Denmark (Project-ID: S-20200114) and Danish Data Agency (Journal no.: 20/49900), the trial was registered at ClinicalTrials.gov (NCT04666584).

### Patient population

Eighty-two patients with stable angina pectoris referred to PCI were enrolled in the study, if they met the inclusion criteria. Patients were eligible if; 1) age was between 18 and 80 years, 2) if they had stable angina pectoris, 3) the target lesion was in a native coronary artery, 4) vessel was suitable for treatment with MgBRS complying with the scaffolds recommended limitations of coronary artery diameter between ≥ 2.75 mm and ≤ 4.0 mm measured with OCT or IVUS. Exclusion criteria were 1) patients participating in other randomized stent studies, 2) expected survival < 1 year, 3) allergy to aspirin, ticagrelor, clopidogrel or prasugrel, 4) allergy to sirolimus, 4) ostial lesions (cannot be cleared with flush by OCT), 5) serum creatinine > 150 µg/L (due to the required amount of contrast by OCT), 6) vastly calcified (evaluated with OCT defined as an arc > 180°, calcium thickness > 0.5 mm and calcium length of > 5 mm), 7) tortuous coronary arteries where the PCI-operator estimated that the introduction of an OCT-catheter would not be possible or would be associated with increased risk, and/or, 8) lesion length > 40 mm. All patients were screened for protocol inclusion and exclusion criteria before enrolment. Patients underwent clinical and invasive imaging follow-up with OCT and IVUS at 6 months.

### Antithrombotic therapy

Patients were treated with aspirin 75 mg /day prior to the PCI procedure. On the day for the PCI, they received a loading dose of 600 mg clopidogrel. Patients were prescribed dual antiplatelet therapy (DAPT) with aspirin 75 mg /day and clopidogrel 75 mg/day for 6 months followed by lifelong monotherapy with 75 mg of aspirin. Patients in Warfarin or novel oral anticoagulant (NOAC) were loaded with 600 mg of clopidogrel. If patients had been admitted and treated for an acute myocardial infraction within the last 12 months, patients kept their previously prescript antithrombotic medication.

### Devices

The metallic-based MgBRS contains a magnesium alloy with a bioresobable poly-L-lactide acid polymer coated with sirolimus as eluting drug released completely after 100 days. The strut thickness is 150 µm. The MgBRS is completely absorbed after 1 year^16^. The scaffold sizes were available in a diameter of 3.0 mm and 3.5 mm, and lengths of 15, 20, and 25 mm.

The scoring balloon (ScoreFlex, OrbusNeich) catheter is a short mono-rail type balloon catheter. It provides forced dilatation with a dual-wire semi-compliant balloon system which facilitates local, safe and controlled plaque modification at lower resolution pressure.

### Procedure strategy

The coronary stenosis was identified by the PCI operator’s interpretation of the angiography and was treated with a MgBRS in all patients. Patients received a dose of heparin (70 UI/kg) prior to the procedure. At the discretion of the operator, pre-dilatation with a 2.0 mm balloon was allowed. Pre-interventional imaging with OCT and IVUS was performed. The scaffold sizing was based on the external elastic membrane (EEM) diameters of the proximal and distal reference segments. If the EEM was visible in >180° of the cross sectional area, the smaller EEM diameter rounded down to the nearest 0.5 mm was used to determine scaffold diameter. If the EEM was visible in <180°, the scaffold diameter was based on the lumen diameter^17^. Patients were allocated 1:1 to either lesion preparation with 1) a scoring balloon, or 2) a standard non-compliant balloon. The lesion was pre-dilated in a 1:1 balloon:artery ratio. Up-scaling to a 0.5 mm larger balloon was allowed, if the pre-dilatation goal was not achieved, as long as the balloon type corresponded to the randomization arm. The pre-dilatation goal was an angiographic residual stenosis of less than 20%. The lesion was then treated with implantation of a MgBRS, and inflation pressure was maintained for 30 seconds during implantation. Mandatory post-dilatation was performed with a non-compliant balloon with the same size or maximally 0.5 mm larger than the implanted scaffold. Lastly, intravascular imaging with OCT and IVUS of the scaffold treated segment was performed and controlled by the PCI-operator and an on-site OCT-analyst. Optimization (if any) was performed at the operators’ discretion. Additional intervention was allowed if there was 1) major under-expansion (minimal scaffold area (MSA) < 4.5 mm^2^), 2) major malapposition (defined as strut > 0.3 mm from the lumen wall for > 3 mm), 3) presence of significant edge dissection, or 4) residual stenosis <5 mm proximal or distal to the scaffold (causing MLA < 4 mm^2^). Repeated OCT and IVUS of the final result were then performed. Blinding of the patient, PCI-operator or investigator to pre-dilatation technique was not possible during the index procedure.

### Intravascular imaging acquisition

OCT and IVUS were performed at baseline and after 6-month of follow-up. The imaging procedures were preceded by administration of 200 µg of intracoronary nitroglycerin. OCT was performed with frequency-domain OPTIS OCT system (Illumien OCT system; Abbott Vascular, Santa Clara, CA, USA) using the Dragonfly^TM^ Imaging catheter. The catheter was positioned 10 mm distally to the lesion or scaffold-treated segment, and the coronary artery was then flushed with 15 ml contrast injection to clear the artery for blood during automated pullback at a rate of 20 mm/s over a distance of 75 mm. The IVUS system (Boston Scientific, Marlborough, MA, USA) used a 40MHz OptiCross 2.6 Fr catheter placed 10 mm distally to the lesion or scaffold-treated segment. Motorized IVUS pullbacks were performed with a pullback speed of 0.5 mm/sec after intracoronary bolus of 200 µg nitroglycerine.

### Intravascular imaging analysis

The intravascular imaging pullbacks were analyzed by two independent analysists who were both blinded to the pre-dilatation technique during analysis. The baseline IVUS and OCT pullbacks were matched with the follow-up images using anatomical landmarks. OCT offline software (Offline Review Workstation; Abbott Vascular) was used for quantitative OCT analysis, and the commercially available program for computerized IVUS-analysis Echoplaque (INDEC Systems, Inc., Santa Clara, CA, USA) was used for IVUS-analysis. The scaffold-treated segment was analyzed for every mm. Lumen dimensions at baseline and follow-up were measured: MLA, mean lumen area, lumen volume, and difference in MLA (follow-up MLA – baseline MLA). Quantitative analysis of scaffold was done using IVUS, because IVUS showed better detection of scaffold remnants than OCT. Scaffold dimensions at baseline were measured: MSA, mean scaffold area, minimum scaffold diameter, and scaffold volume. Scaffold malapposition was defined to be present when the distance between the abluminal surface of the strut and the luminal surface of the vessel wall exceeded the struts thickness of 150 µm. Major malapposition was defined as struts > 0.3 mm from the lumen wall for >3 mm in length^18^, and the remaining were classified as minor. At baseline, malapposition area, distance, and volume were analyzed. At follow-up, visible struts or strut remnants were categorized as malapposed when the abluminal border of the strut/remnant was separated from the lumen surface by a visible space exceeding 150 µm. The malapposition observations was matched from baseline to follow-up and divided into resolved, persistent, or late acquired malapposition. If a scaffold contained both resolved and persistent malapposition at follow-up, it was summarized as persistent. To evaluate the effect of pre-dilatation method on remodeling in the specific lesion site, IVUS was used to identify the pre-procedure MLA in lesion. The lesion site was defined as 5 mm proximally and distally to MLA. The corresponding 10 mm segment was identified in IVUS pullback post-procedure and at 6-month follow-up using anatomical landmarks such as side branches, calcified plaques and scaffold edges. Remodeling was defined as changes in mean EEM area in the lesion site and deemed significant if the mean EEM area changed more than 0.5 mm^2^. Enlargement was defined as positive remodeling, and reduction in mean EEM area was defined as negative remodeling. Quantitative IVUS analysis included measurements of EEM, peri-scaffold plaque (EEM area – scaffold area), and total plaque area (EEM area – lumen area).

## Statistical analysis

Categorical data was presented as numbers and frequencies and compared using chi-square test or Fisher’s exact statistics. Continuous data was presented as mean ± SD and compared using Student’s t-test. Paired t-test was used for comparison from baseline to follow-up. If the distribution were skewed, a non-parametric test was performed, and median with interquartile range (IQR) was stated.

All tests were two-tailed, and a p-value <0.05 was considered statistically significant. STATA version 18.0 (StataCorp, Collage Station, TX, USA) was used for the statistical analysis. Inter-observer variability for imaging analysis was tested for consistency of agreement using an intraclass correlation coefficient (ICC) was calculated for MLA at follow-up and for malapposition area at baseline and follow-up. The Pearson correlation coefficient was used to evaluate the direction and strength of the linear relation between two parameters.

The estimated sample size was based on data from the HONEST study^19^. The reduction of MLA from 6.99 mm^2^ to 5.01 mm^2^ (27%) 6 months after implantation of the Magmaris BVS, represented the expected reference group. Optimal lesion preparation with pre-dilatation with a scoring balloon is estimated to minimize MLA reduction from 6.99 mm^2^ to 6.22 mm^2^ (11%). A power calculation is conducted using the expected MLA after 6 months (6.22 mm^2^ for the scoring balloon and 5.01 mm^2^ for the standard non-compliant balloon). Inclusion of 35 patients in each group is necessary to reach statistical significance in case of 2-tailed significance level of 0.05 and power of 80 %. Loss to follow-up and poor image quality finalize an expected drop-out rate of 15 %, thereby requiring 82 patients in total.

## Endpoints

The primary endpoint was MLA in the scaffold-treated segment pre-dilated with a scoring balloon versus standard non-compliant balloon 6-month after implantation of a MgBRS assessed with OCT.

Secondary endpoints were differences between treatment groups in: 1) change in MLA, and 2) percentage and size of incomplete scaffold apposition at baseline and follow-up.

## Results

A flowchart of enrolled patients is provided in Figure 1.

**Figure 1:**
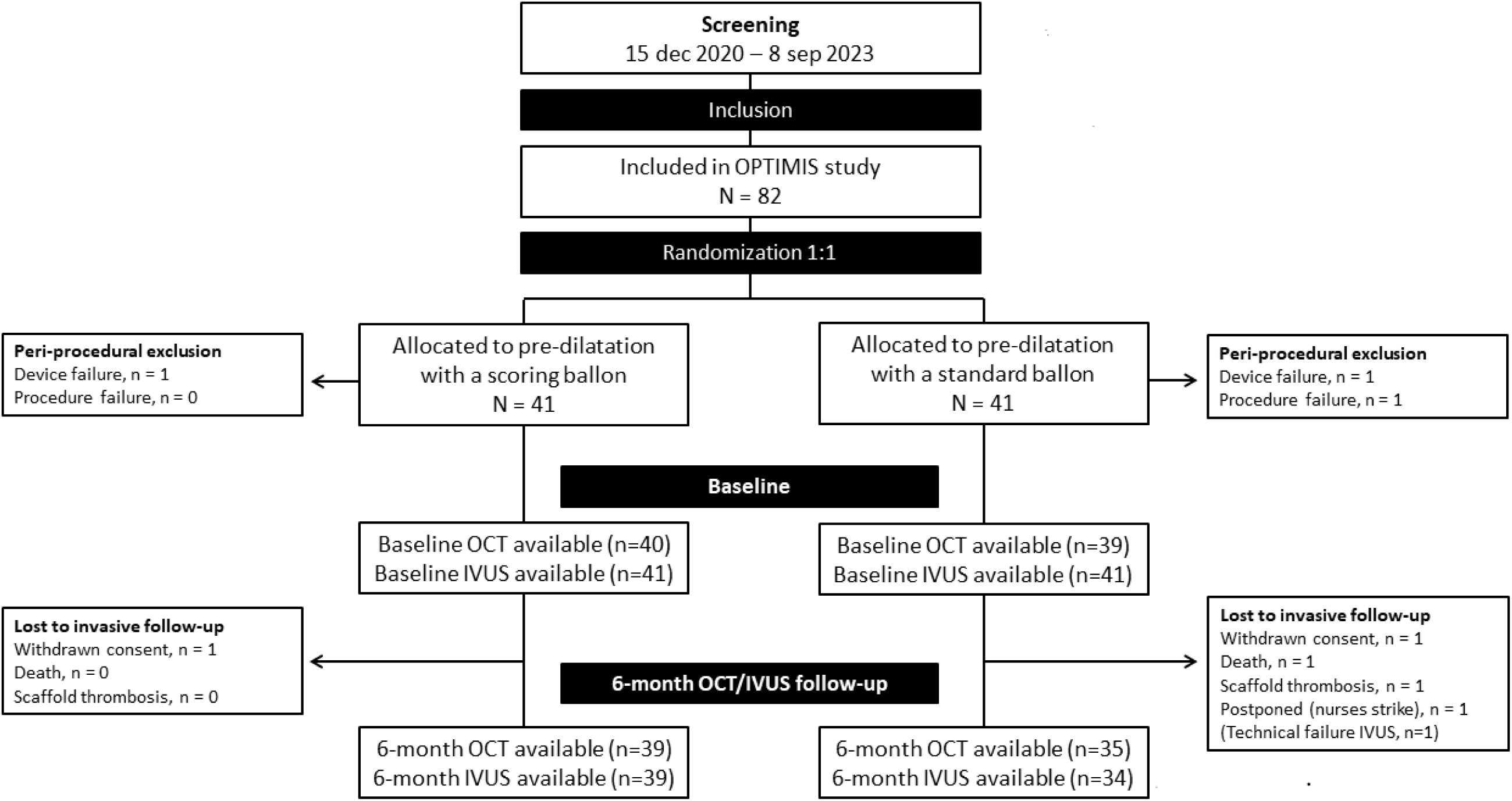
Flow chart

In total, 82 patients were enrolled in the study. Follow-up images were not available in 8 patients due to following reasons. One patient randomized to standard non-compliant balloon pre-dilatation was excluded due to vessel dissection that could not be covered by a MgBRS scaffold. Two patients were excluded, one in the scoring balloon group and one in the standard non-compliant balloon group, due to scaffold failure where the MgBRS was lost in the coronary artery proximally to the study lesion. In all three cases, patients were treated with a DES. Five patients had unavailable follow-up images: Two patients withdrew consents (one in the scoring balloon group and one standard non-compliant balloon group), one patient died within the 6-month angiographic follow-up (standard non-compliant balloon group), one patient had a subacute scaffold thrombosis 5 days after implantation (standard non-compliant balloon group), and one patient was postponed due to nurses’ strike (standard non-compliant balloon group).

### Clinical and procedural characteristics

Baseline clinical and procedural characteristics are presented in Table 1 and Table 2.

**Table 1:**
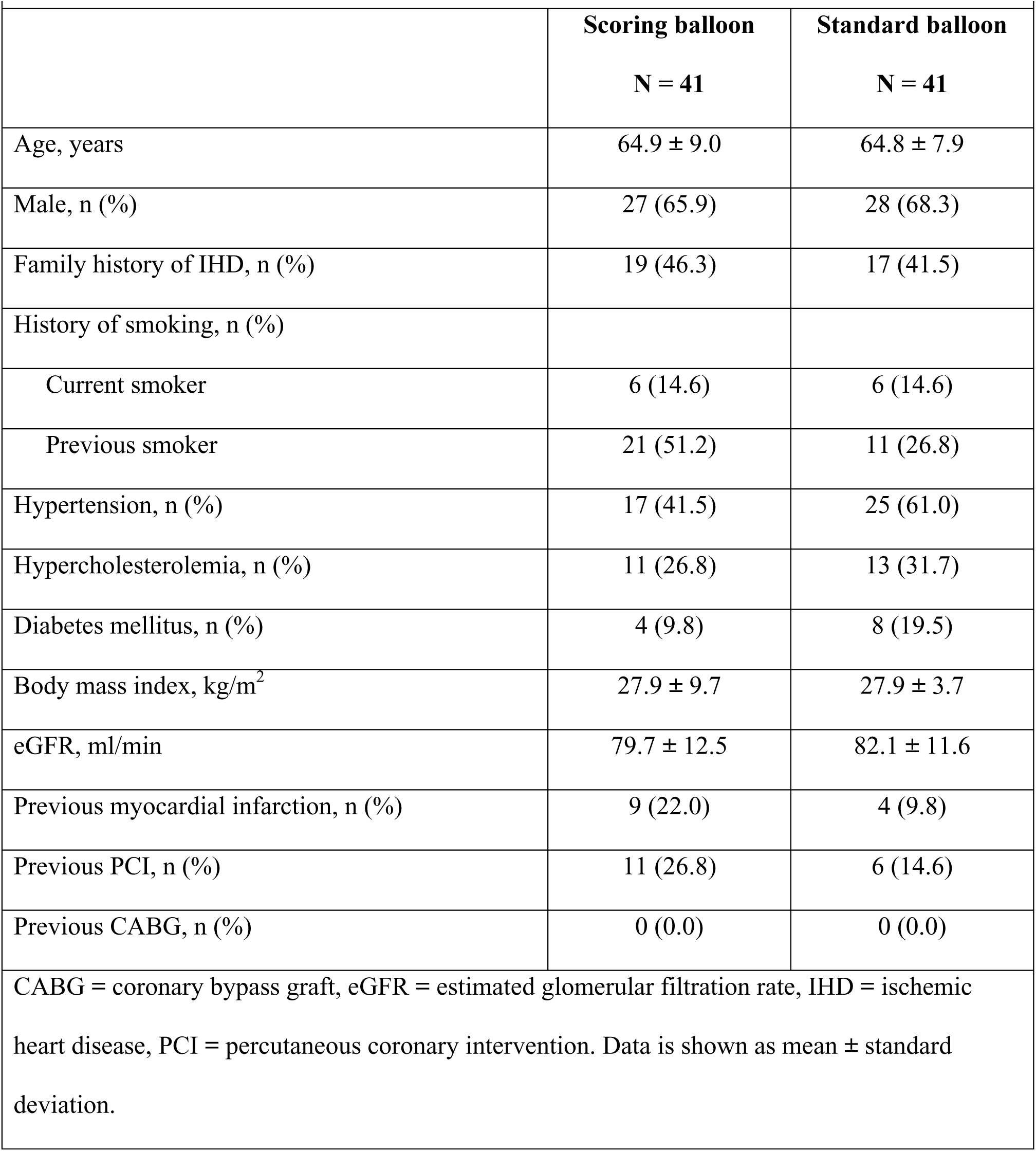
Patient baseline characteristics.

| **Table 1: Patient baseline characteristics** |  |  |
| --- | --- | --- |
|  | **Scoring balloon** | **Standard balloon** |
|  | **N = 41** | **N = 41** |
| Age, years | 64.9 ± 9.0 | 64.8 ± 7.9 |
| Male, n (%) | 27 (65.9) | 28 (68.3) |
| Family history of IHD, n (%) | 19 (46.3) | 17 (41.5) |
| History of smoking, n (%) |  |  |
| Current smoker | 6 (14.6) | 6 (14.6) |
| Previous smoker | 21 (51.2) | 11 (26.8) |
| Hypertension, n (%) | 17 (41.5) | 25 (61.0) |
| Hypercholesterolemia, n (%) | 11 (26.8) | 13 (31.7) |
| Diabetes mellitus, n (%) | 4 (9.8) | 8 (19.5) |
| Body mass index, kg/m<sup>2</sup> | 27.9 ± 9.7 | 27.9 ± 3.7 |
| eGFR, ml/min | 79.7 ± 12.5 | 82.1 ± 11.6 |
| Previous myocardial infarction, n (%) | 9 (22.0) | 4 (9.8) |
| Previous PCI, n (%) | 11 (26.8) | 6 (14.6) |
| Previous CABG, n (%) | 0 (0.0) | 0 (0.0) |
| CABG = coronary bypass graft, eGFR = estimated glomerular filtration rate, IHD = ischemic heart disease, PCI = percutaneous coronary intervention. Data is shown as mean ± standard deviation. |  |  |

**Table 2:** Procedural and angiographic characteristics.

| **Table 2: Procedural and angiographic characteristics** |  |  |
| --- | --- | --- |
|  | Scoring balloon<br>N = 41 | Standard balloon<br>N = 41 |
| Target coronary artery, n (%) |  |  |
| Left anterior descending | 23 (56.1) | 24 (58.5) |
| Left circumflex | 6 (14.6) | 8 (19.5) |
| Right coronary artery | 12 (29.3) | 9 (22.0) |
| Lesion length, mm | 23.9 ± 10.5 | 22.5 ± 5.4 |
| Reference vessel diameter, mm | 3.4 ± 0.3 | 3.4 ± 0.3 |
| Pre-dilatation, n (%) | 41 (100) | 41 (100) |
| Balloon diameter at pre-dilatation, mm | 3.3 ± 0.3 | 3.3 ± 0.3 |
| Balloon length at pre-dilatation, mm | 13.1 ± 2.5* | 15.5 ± 3.3 |
| Max balloon pressure at pre-dilatation, atm | 13.1 ± 2.8 | 14.0 ± 2.7 |
| Number of scaffolds per lesion, mm | 1.1 ± 0.3 | 1.1 ± 0.2 |
| Scaffold length, mm | 19.4 ± 4.1 | 21.0 ± 3.9 |
| Scaffold diameter, mm | 3.3 ± 0.2 | 3.3 ± 0.3 |
| Maximum balloon pressure, atm | 11.9 ± 2.2 | 12.0 ± 2.4 |
| Expected scaffold diameter, mm | 3.4 ± 0.3 | 3.4 ± 0.3 |
| Post-dilatation, n (%) | 38 (93) | 41 (100) |
| Balloon diameter at post-dilatation, mm | 3.7 ± 0.3 | 3.7 ± 0.4 |
| Balloon length at post-dilatation, mm | 14.5 ± 3.6 | 15.7 ± 3.3 |
| Max balloon pressure at post-dilatation, atm | 13.0 ± 2.6 | 12.8 ± 2.4 |
| Flouro time, minutes | 12.3 ± 5.3 | 11.8 ± 5.2 |
| Contrast volume, ml | 108.0 ± 42.9 | 102.6 ± 43.8 |
| Procedure time, minutes | 49.9 ± 17.7 | 47.9 ± 18.4 |
| * significantly shorter balloon length at pre-dilatation in the scoring balloon group (p-value < 0.001) |  |  |

The treatment groups were well matched without any significant differences in baseline characteristics. Also, there were no significant differences in procedural characteristics, except for balloon length which was significantly shorter in the scoring balloon group (only available in 10 and 15 mm) (13.1 ± 2.5 mm vs. 15.5 ± 3.3 mm, p < 0.001) compared to the standard non-compliant balloon group.

### Optical coherence tomography findings

Post-procedure and 6-month follow-up OCT findings are presented in Table 3. Inter-observer variability for MLA at follow-up was: ICC=0.996 (95% confidence interval (CI): 0.999-1.00, p<0.001), for total malapposition area at baseline: ICC=0.949 (95% CI: 0.77-0.99, p< 0.001), and for total malapposition at follow-up: ICC=0.874 (95% CI: 0.50-0.97, p=0.001).

**Table 3:**
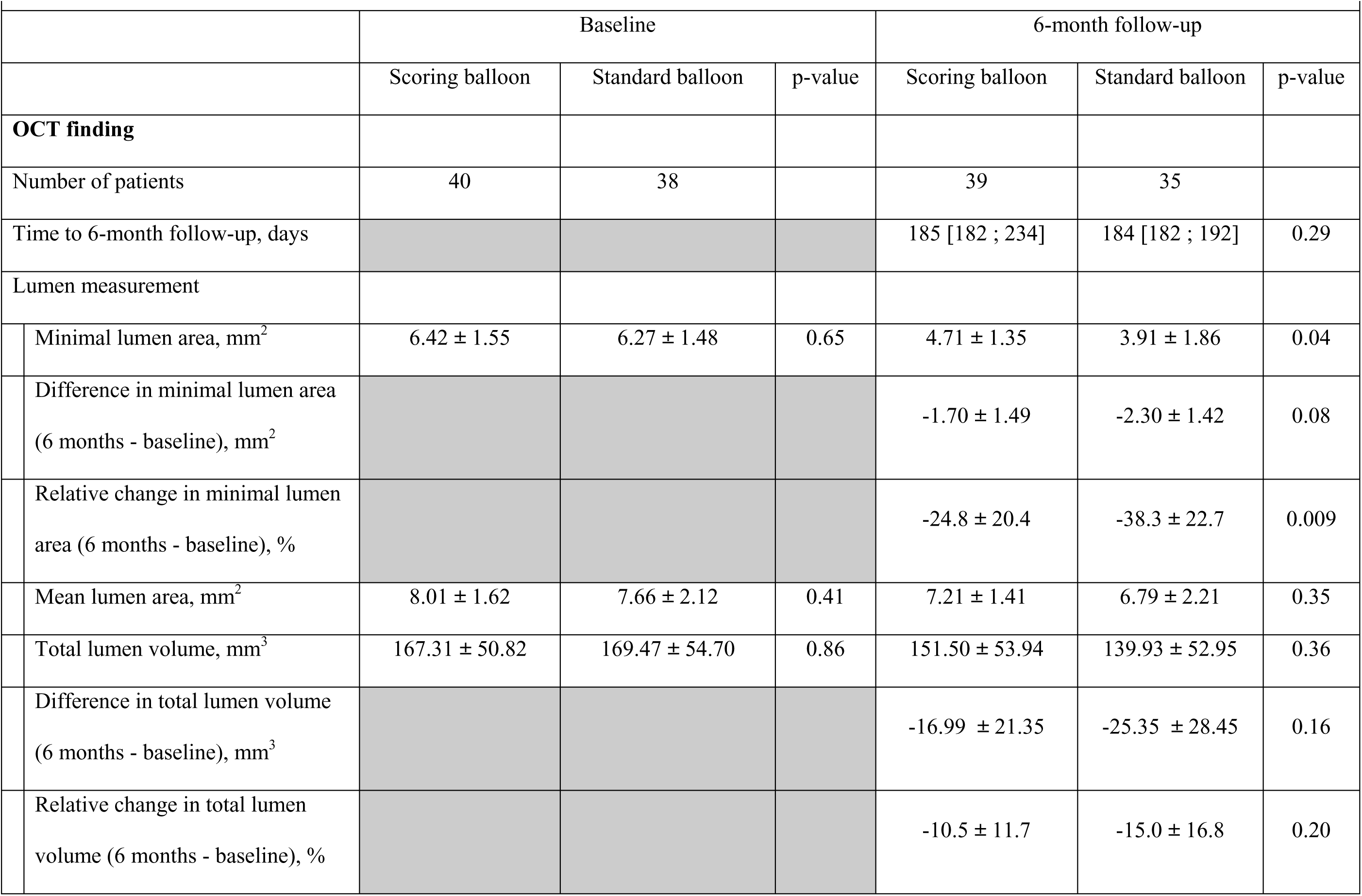
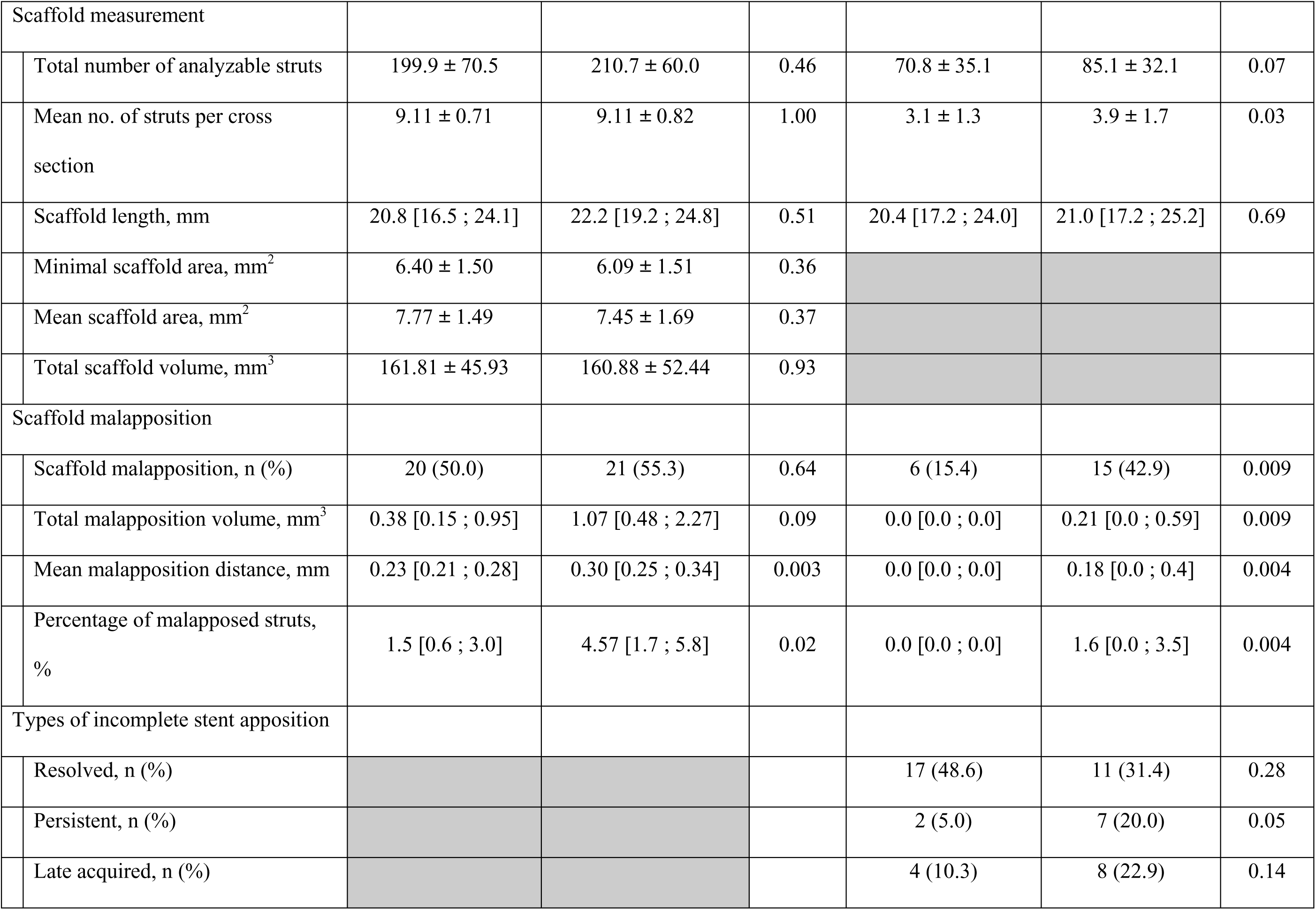
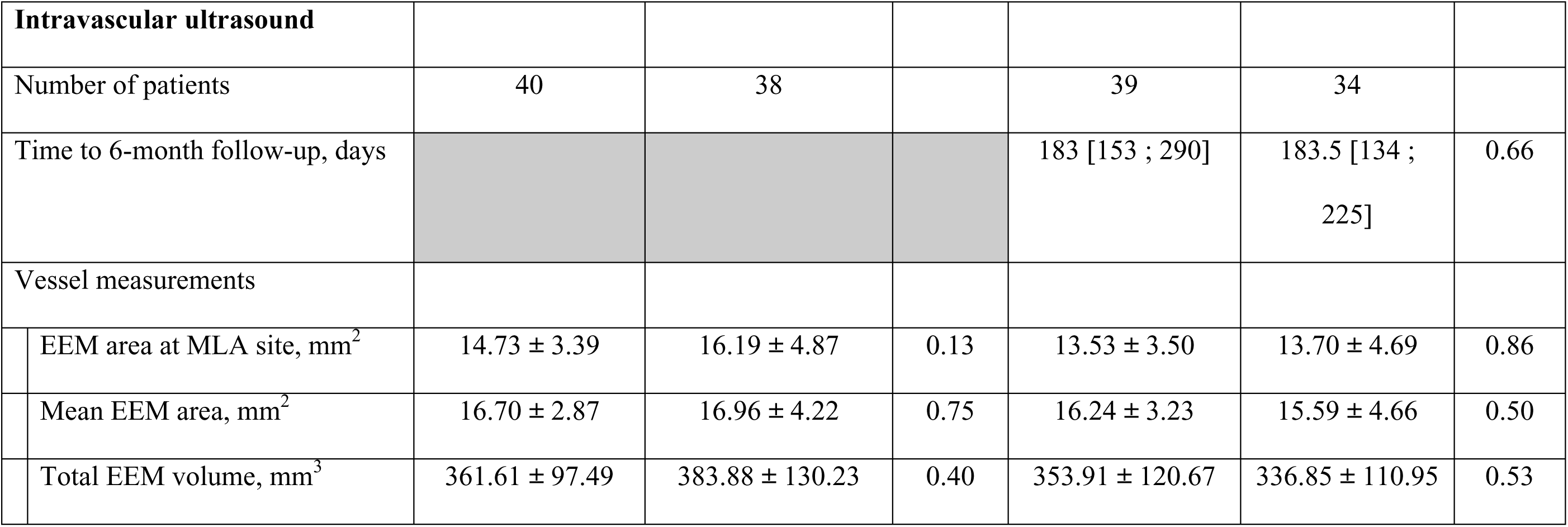
Post-procedure and 6-month follow-up optical coherence tomography findings and intravascular ultrasound.

|  |  |  |  |  |  |  |
| --- | --- | --- | --- | --- | --- | --- |
|  | Baseline |  |  | 6-month follow-up |  |  |
|  | Scoring balloon | Standard balloon | p-value | Scoring balloon | Standard balloon | p-value |
| **OCT finding** |  |  |  |  |  |  |
| Number of patients | 40 | 38 |  | 39 | 35 |  |
| Time to 6-month follow-up, days |  |  |  | 185 [182 ; 234] | 184 [182 ; 192] | 0.29 |
| Lumen measurement |  |  |  |  |  |  |
| Minimal lumen area, mm<sup>2</sup> | 6.42 ± 1.55 | 6.27 ± 1.48 | 0.65 | 4.71 ± 1.35 | 3.91 ± 1.86 | 0.04 |
| Difference in minimal lumen area<br>(6 months - baseline), mm<sup>2</sup> |  |  |  | -1.70 ± 1.49 | -2.30 ± 1.42 | 0.08 |
| Relative change in minimal lumen<br>area (6 months - baseline), % |  |  |  | -24.8 ± 20.4 | -38.3 ± 22.7 | 0.009 |
| Mean lumen area, mm<sup>2</sup> | 8.01 ± 1.62 | 7.66 ± 2.12 | 0.41 | 7.21 ± 1.41 | 6.79 ± 2.21 | 0.35 |
| Total lumen volume, mm<sup>3</sup> | 167.31 ± 50.82 | 169.47 ± 54.70 | 0.86 | 151.50 ± 53.94 | 139.93 ± 52.95 | 0.36 |
| Difference in total lumen volume<br>(6 months - baseline), mm<sup>3</sup> |  |  |  | -16.99 ± 21.35 | -25.35 ± 28.45 | 0.16 |
| Relative change in total lumen<br>volume (6 months - baseline), % |  |  |  | -10.5 ± 11.7 | -15.0 ± 16.8 | 0.20 |

|  |  |  |  |  |  |  |  |
| --- | --- | --- | --- | --- | --- | --- | --- |
| Scaffold measurement |  |  |  |  |  |  |  |
|  | Total number of analyzable struts | 199.9 ± 70.5 | 210.7 ± 60.0 | 0.46 | 70.8 ± 35.1 | 85.1 ± 32.1 | 0.07 |
|  | Mean no. of struts per cross section | 9.11 ± 0.71 | 9.11 ± 0.82 | 1.00 | 3.1 ± 1.3 | 3.9 ± 1.7 | 0.03 |
|  | Scaffold length, mm | 20.8 [16.5 ; 24.1] | 22.2 [19.2 ; 24.8] | 0.51 | 20.4 [17.2 ; 24.0] | 21.0 [17.2 ; 25.2] | 0.69 |
|  | Minimal scaffold area, mm<sup>2</sup> | 6.40 ± 1.50 | 6.09 ± 1.51 | 0.36 |  |  |  |
|  | Mean scaffold area, mm<sup>2</sup> | 7.77 ± 1.49 | 7.45 ± 1.69 | 0.37 |  |  |  |
|  | Total scaffold volume, mm<sup>3</sup> | 161.81 ± 45.93 | 160.88 ± 52.44 | 0.93 |  |  |  |
| Scaffold malapposition |  |  |  |  |  |  |  |
|  | Scaffold malapposition, n (%) | 20 (50.0) | 21 (55.3) | 0.64 | 6 (15.4) | 15 (42.9) | 0.009 |
|  | Total malapposition volume, mm<sup>3</sup> | 0.38 [0.15 ; 0.95] | 1.07 [0.48 ; 2.27] | 0.09 | 0.0 [0.0 ; 0.0] | 0.21 [0.0 ; 0.59] | 0.009 |
|  | Mean malapposition distance, mm | 0.23 [0.21 ; 0.28] | 0.30 [0.25 ; 0.34] | 0.003 | 0.0 [0.0 ; 0.0] | 0.18 [0.0 ; 0.4] | 0.004 |
|  | Percentage of malapposed struts, % | 1.5 [0.6 ; 3.0] | 4.57 [1.7 ; 5.8] | 0.02 | 0.0 [0.0 ; 0.0] | 1.6 [0.0 ; 3.5] | 0.004 |
| Types of incomplete stent apposition |  |  |  |  |  |  |  |
|  | Resolved, n (%) |  |  |  | 17 (48.6) | 11 (31.4) | 0.28 |
|  | Persistent, n (%) |  |  |  | 2 (5.0) | 7 (20.0) | 0.05 |
|  | Late acquired, n (%) |  |  |  | 4 (10.3) | 8 (22.9) | 0.14 |
| **Intravascular ultrasound** |  |  |  |  |  |  |  |
| Number of patients |  | 40 | 38 |  | 39 | 34 |  |
| Time to 6-month follow-up, days |  |  |  |  | 183 [153 ; 290] | 183.5 [134 ; 225] | 0.66 |
| Vessel measurements |  |  |  |  |  |  |  |
|  | EEM area at MLA site, mm<sup>2</sup> | 14.73 ± 3.39 | 16.19 ± 4.87 | 0.13 | 13.53 ± 3.50 | 13.70 ± 4.69 | 0.86 |
|  | Mean EEM area, mm<sup>2</sup> | 16.70 ± 2.87 | 16.96 ± 4.22 | 0.75 | 16.24 ± 3.23 | 15.59 ± 4.66 | 0.50 |
|  | Total EEM volume, mm<sup>3</sup> | 361.61 ± 97.49 | 383.88 ± 130.23 | 0.40 | 353.91 ± 120.67 | 336.85 ± 110.95 | 0.53 |

#### Lumen dimensions

At baseline, there was no significant difference in MLA, mean LA, or lumen volume between the two treatment groups assessed with OCT. At 6-month follow-up, MLA (the primary endpoint) in the scaffold-treated segment was significantly larger in the patients allocated to pre-dilatation with a scoring balloon, compared to a standard non-compliant balloon (4.71 mm^2^ ± 1.35 vs. 3.91 mm^2^ ± 1.86, p = 0.04). There was no significant difference between the two groups in mean LA, or lumen volume at 6-month follow-up. There was a relative reduction in MLA of -24.8% for the scoring balloon group compared to -38.3% in the standard non-compliant balloon group, p=0.009. Representative cases of lumen reduction from baseline to follow-up are shown in Figure 2.

**Figure 2:**
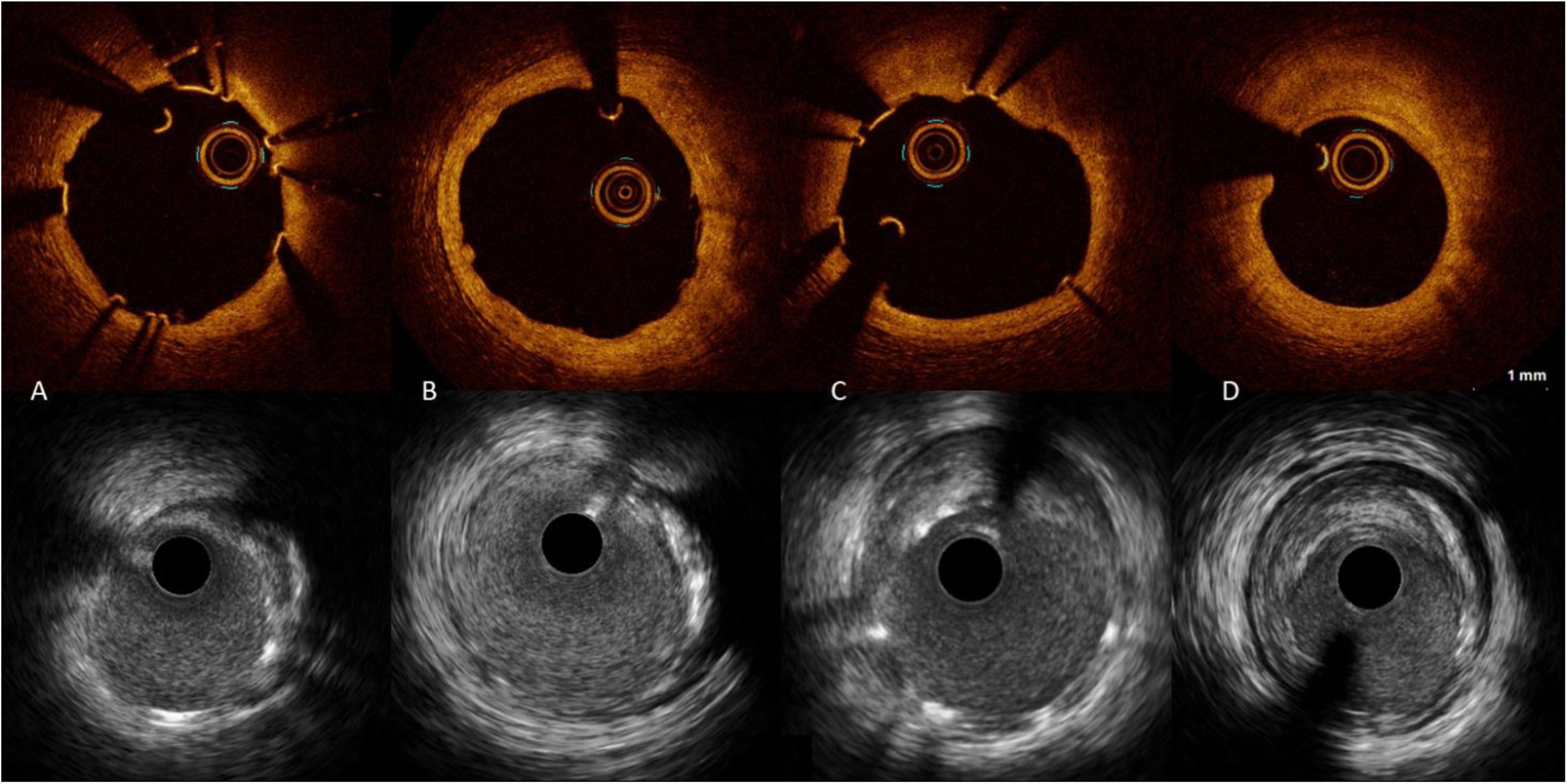
Intravascular images of lumen reduction after implantation of Magmaris bioresorbable scaffold. The upper panel shows OCT images of minimal lumen area from baseline and the corresponding site at follow-up. The lower panel shows the matching site acquired with IVUS. Images A and B represent the vascular healing after lesion preparation with a scoring balloon prior to MgBRS implantation. Lumen area at baseline was 7.3 mm^2^ measured with OCT and 7.5 mm^2^ with IVUS. Vessel area was 12.7 mm^2^ at baseline (A). At 6-month follow-up, lumen area was 8.8 mm^2^ with OCT and 8.8 mm^2^ with IVUS. Vessel area was 16.0 mm^2^ (B). Images C and D represent the vascular healing after implantation of a MgBRS in a lesion pre-dilated with a standard non-compliant balloon. Lumen area at baseline was 8.8 mm^2^ with OCT and 8.8 mm^2^ with IVUS. Vessel area was 16.0 mm^2^ (C). After 6 months, the matching site was reduced to 5.1 mm^2^ measured with OCT, and 5.6 mm^2^ with IVUS. Vessel area was 13.8 mm^2^ (D). Abbreviations: IVUS = Intravascular ultrasound; MgBRS = Magmaris bioresorbable scaffold; OCT = Optical coherence tomography.

#### Scaffold measurements and malapposition

At baseline, scaffold parameters, such as scaffold length, mean scaffold area, minimal scaffold area, and total scaffold volume were similar in the two groups. Total number of analyzable struts were similar at baseline between the two groups (199.9 ± 70.5 in the scoring balloon group and 210.7 ± 60.0 in the standard non-compliant balloon group, p=0.46). At follow-up, the total number of analyzable struts were reduced to 70.8 ± 35.1 in the scoring balloon group and 85.1 ± 32.1 in the standard non-compliant balloon group (p=0.07).

At baseline, half of the scaffolds in both groups had minor malapposition. There were no major malappositions in any of the groups. Percentage of malapposed struts was small in both groups and significantly lower in the scoring balloon group with 1.5 % compared to 4.6 % in the standard non-compliant balloon group (p=0.02). At baseline, malapposition volume tended to be smaller in the scoring balloon group (0.38 mm^2^ [0.15 ; 0.95]) compared to the standard non-compliant balloon group [1.07 mm^2^ 0.48 ; 2.27], but there was no significant difference (p= 0.09).

At 6-month follow-up, 15.4% of the lesions treated with the scoring balloon had minor malappositions, whereas 42.9% in the standard balloon group had minor malappositions (p=0.009). There significantly smaller total malapposition volume (0.0 [0.0 ; 0.0] vs. 0.21 [0.0 ; 0.59], p=0.009) and percentage of malapposed struts (0.0 [0.0 ; 0.0] vs. 1.62 [0.0 ; 3.49], p=0.004) in the scoring group compared to the standard non-compliant balloon group at 6-month follow-up. Type of malapposition did not differ between groups. Malappositions were resolved in 31.4 % of the scaffolds in the scoring balloon group, compared to 48.6% in the standard non-compliant balloon group. In the scoring balloon group, 5% had persistent malapposition vs. 20% in the standard balloon group. Late acquired malapposition was seen in 15.4% in the scoring balloon group compared to 22.9% in the standard non-compliant balloon group, and often positioned at scaffold edge and in relation to calcified plaque. Malapposition types are presented in Figure 3.

**Figure 3:**
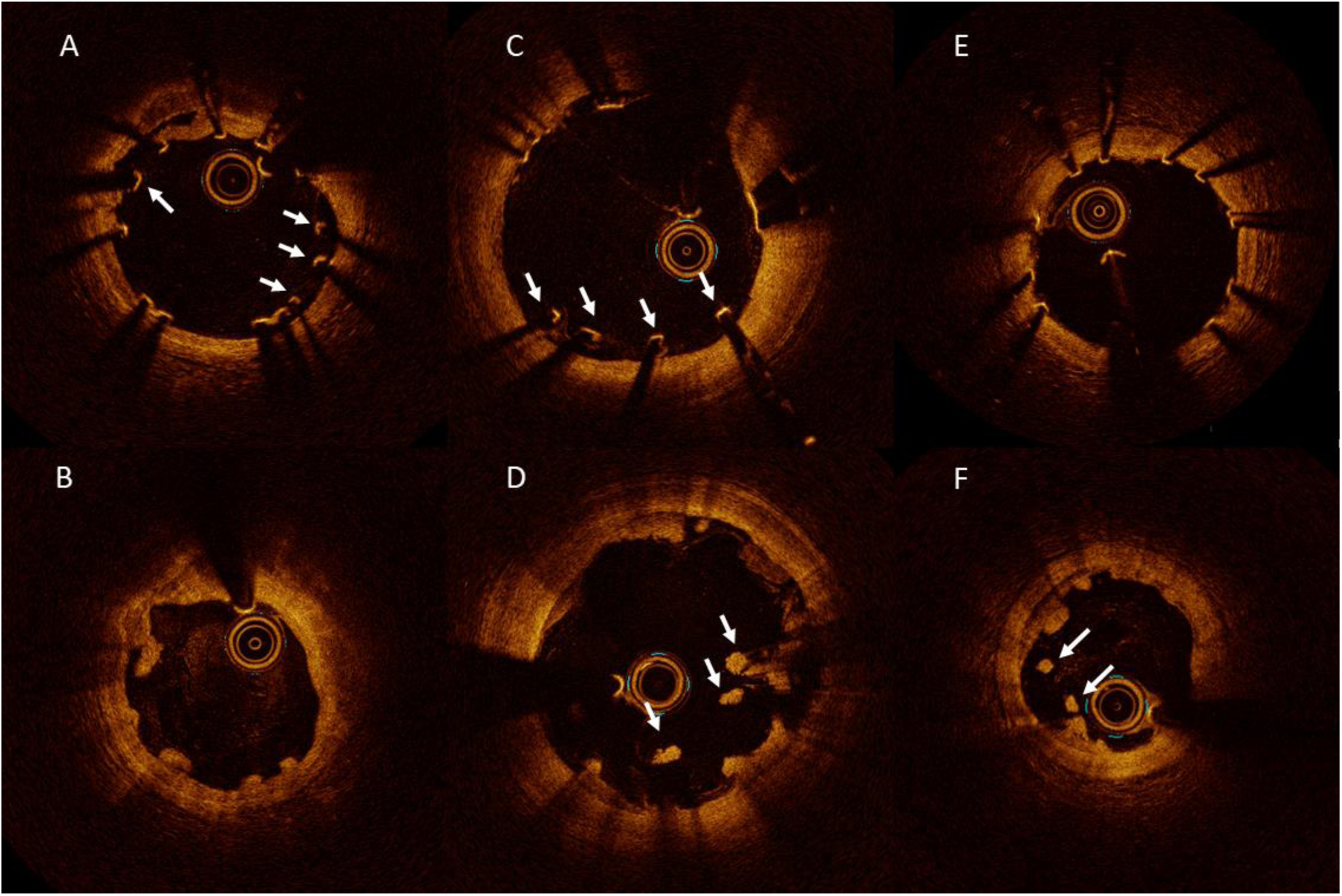
Optical coherence tomography images of malapposition types Upper panel represents baseline optical coherence tomography images, and lower panel represents 6-month follow-up. A) There are three malapposed struts from 3 to 5 o’clock, and one malapposed strut at 10 o’clock. The corresponding site after 6 months (B) revealed resolved malapposition from 3 to 5 o’clock, but persistent malapposition at 10 o’clock. C) Four malapposed struts are visible at baseline from 5 to 7 o’clock. At 6-month follow-up, persistent malapposition is seen in the corresponding cross section (D). E) All struts are well-apposed, but after 6 month acquired malapposition appears at 7 to 8 o’clock.

At 6-month follow-up, no scaffold area and volume were drawn since most of the struts were absorbed. OCT images of scaffold degradation are shown in Figure 2. The total number of struts were similar in the two groups, but there were significantly less struts per cross section in the scoring balloon group compared to standard non-compliant balloon group after 6 months.

### Intravascular ultrasound findings

Post-procedure and 6-month follow-up IVUS findings are presented in Table 3 and Table 4 and Supplementary table 1.

**Table 4:** Remodeling of lesion segment pre-procedure and corresponding segment post-procedure and at 6-month follow-up assessed with IVUS.

|  |  |  |  |
| --- | --- | --- | --- |
|  | Scoring<br>N = 39 | Standard<br>N = 34 | p-value |
| Mean lumen (mm<sup>2</sup>) |  |  |  |
| Pre-procedure | 5.3 ± 1.4 | 4.8 ± 1.5 | 0.13 |
| Post-procedure | 8.5 ± 1.4 | 8.2 ± 1.7 | 0.31 |
| 6-month follow-up | 8.1 ± 1.8 | 7.4 ± 2.6 | 0.19 |
| Change (6 months – baseline) | -0.4 ± 1.5 | -0.8 ± 1.6 | 0.41 |
| p-value (baseline vs. 6 months)* | 0.08 | 0.009 |  |
| Mean EEM area (mm<sup>2</sup>) |  |  |  |
| Pre-procedure | 13.3 ± 3.1 | 13.4 ± 4.8 | 0.88 |
| Post-procedure | 16.8 ± 2.9 | 17.1 ± 4.4 | 0.74 |
| 6-month follow-up | 17.0 ± 3.6 | 15.7 ± 4.9 | 0.20 |
| Change (6 months – baseline) | 0.2 ± 2.0 | -1.4 ± 2.0 | 0.001 |
| p-value (baseline vs. 6 months)* | 0.62 | < 0.001 |  |
| Mean plaque area (mm<sup>2</sup>) |  |  |  |
| Pre-procedure | 7.9 ± 2.5 | 8.6 ± 3.9 | 0.39 |
| Post-procedure | 8.3 ± 2.0 | 8.9 ± 3.5 | 0.31 |
| 6-month follow-up | 8.9 ± 2.4 | 8.3 ± 3.0 | 0.36 |
| Change (6 months – baseline) | 0.6 ± 1.3 | -0.7 ± 1.9 | 0.002 |
| p-value (baseline vs. 6 months)* | 0.007 | 0.06 |  |
| Mean scaffold area (mm<sup>2</sup>) |  |  |  |
| Post-procedure | $9.5 \pm 1.7$ | $9.1 \pm 1.9$ | 0.26 |
| 6-month follow-up | $10.1 \pm 2.1$ | $8.9 \pm 2.8$ | 0.04 |
| Change (6 months – baseline) | $0.6 \pm 1.9$ | $-0.2 \pm 1.8$ | 0.10 |
| p-value (baseline vs. 6 months)* | 0.07 | 0.58 |  |
| *paired analysis |  |  |  |

#### Vessel dimensions

There was no difference in vessel measurements between the two groups at baseline or at 6-month follow-up (Table 3). The paired analysis of mean area in the 10 mm lesion site and corresponding segment post-procedure and at 6-month follow-up are presented in Table 4. There was no significant difference in mean lumen area from post-procedure to 6-month follow-up in the scoring balloon group (8.5 ± 1.4 mm^2^ vs. 8.1 ± 1.8 mm^2^, p=0.08), whereas a significant decrease in lumen area was found in the standard non-compliant balloon group (8.2 ± 1.7 mm^2^ vs. 7.4 ± 2.6 mm^2^, p=0.009). Vessel area in the 10 mm segment corresponding to the lesion site did not change in the scoring balloon group from baseline to 6-month follow-up (16.8 ± 2.9 mm^2^ vs. 17.0 ± 3.6 mm^2^, p=0.62), but was significantly decreased (17.1 ± 4.4 mm^2^ vs. 15.7 ± 4.9 mm^2^, p < 0.001) in the standard non-compliant balloon group indicating negative remodeling.

#### Pattern of remodeling

Figure 4 shows the relationship between relative change in lumen area and relative change in vessel area (A), and relative change in lumen area and relative change in plaque area (B). There was a significant positive correlation between relative change in lumen area and relative change in vessel area at the 10 mm lesion site (r=0.72, 95% CI: 0.58-0.81, p<0.001), but there was no correlation between relative change in lumen area and relative change in plaque area (r=-0.02, 95% CI: -0.25-0.21, p=0.88).

**Figure 4:**
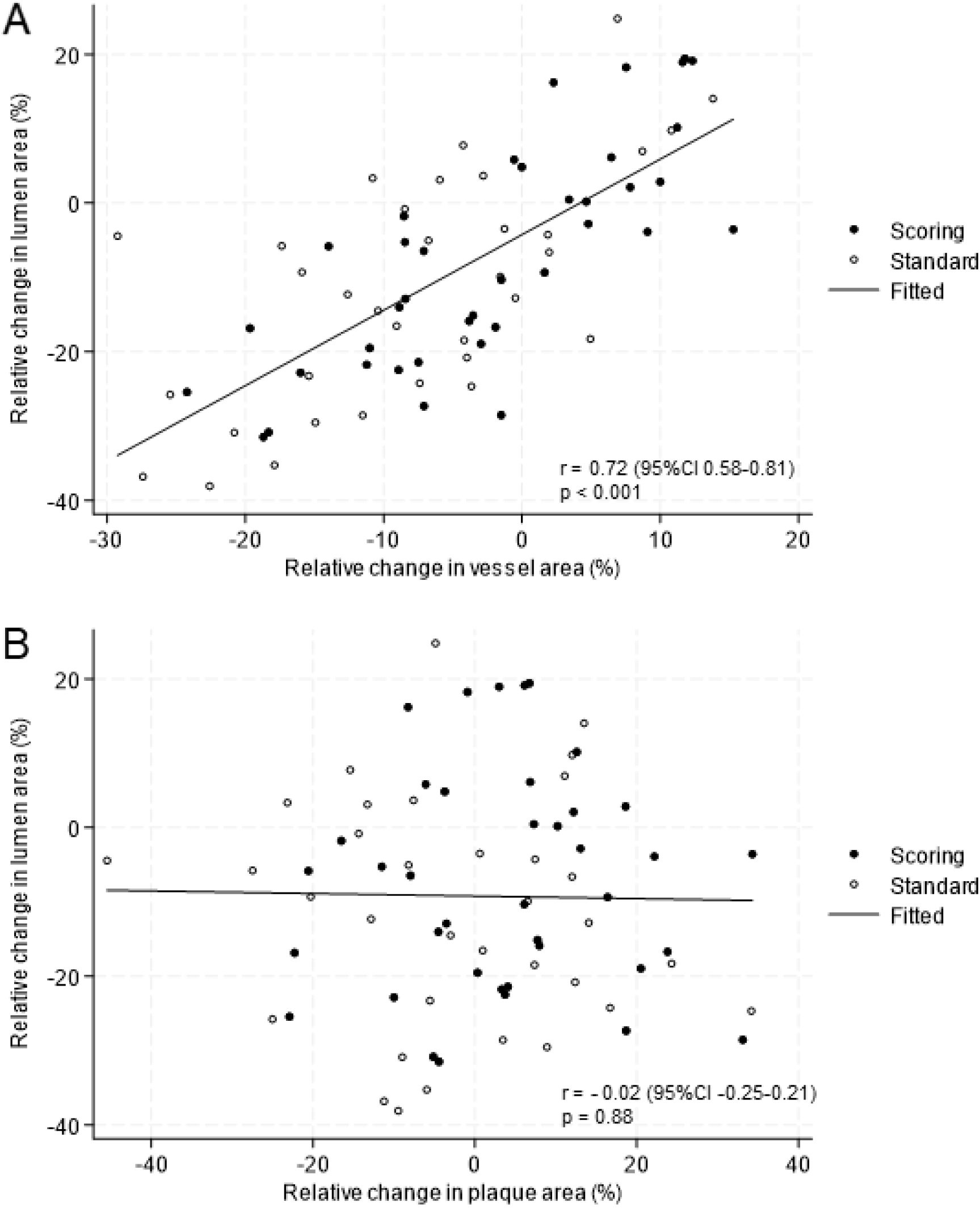
Pattern of remodeling at the lesion site A. Correlation between relative change in lumen area (%) and relative change in vessel area (%) at the lesion site. B. Correlation between relative change in lumen area (%) and relative change in plaque area at the lesion site

### Clinical 6-month follow-up

In patients allocated to pre-dilatation with a scoring balloon prior to implantation of the MgBRS one patient had a target vessel revascularization not related to the scaffold-treated segment. There were no events observed corresponding to the scaffold-treated segment in the scoring balloon group. In patients treated with the standard balloon prior to implantation of the MgBRS following events were observed: one patient admitted with STEMI and subacute scaffold thrombosis 5 days after index procedure. This patient was only treated with aspirin for 4 days followed by monotherapy with clopidogrel as the patient also received NOAC; one patient died due to an intracranial hemorrhage 92 days after index procedure.

## Discussion

In summary, we found that MLA assessed with OCT was significantly larger in the scoring balloon group compared to the standard non-compliant balloon group 6 month after implantation of the MgBRS. In both groups, MLA decreased from baseline to 6-month follow-up, but less MLA reduction was seen in the scoring balloon group compared to the standard non-compliant balloon group. At the lesion site, there was no change in remodeling from baseline to follow-up in the scoring balloon group, whereas negative remodeling was observed in lesions prepared with the standard non-compliant balloon. In the lesions pre-dilated with a scoring balloon, there was significantly less malapposition at follow-up compared to the standard non-compliant balloon group.

The magnesium-based BRS was first evaluated in the DREAM 1G study^20^, where a significant decrease in MLA was observed within the first 6 months (7.9 mm^2^ ± 1.2 vs. 5.7 mm^2^ ± 1.0) after implantation assessed with OCT. The second generation magnesium-based BRS, MgBRS, had higher flexibility and higher radial force, than the first generation magnesium-based BRS^21^.

Previous studies have investigated the vascular healing after 6 months of the magnesium-based BRS with both IVUS and OCT, but significant lumen decrease continued to occur^6, 19, 22, 23^.

Assessed with OCT, malapposition, neointimal hyperplasia and strut coverage were near impossible to detect at follow-up, because the strut remnants had lost their metallic stent-like appearance during the absorption process. Interestingly, the BIOSOLVE-II study (BIOtroniks – Safety and performance in de nOvo Lesion of natiVE coronary arteries with Magmaris) reported measurable scaffold observation, such as mean and minimum scaffold area and incomplete strut apposition as visible with IVUS, but not with OCT at 6-month follow-up^22^. The same pattern applied to our findings, where scaffold area detection was not possible with OCT, but analyzable with IVUS at 6-month follow-up. The BIOSOLVE-II trial^22^ measured smaller lumen and scaffold areas assessed with IVUS compared to OCT, which was unlike our findings with smaller lumen and scaffold measurements evaluated with OCT compared to IVUS. IVUS is often reported to overestimate lumen area compared to OCT^17^, which may explain why no difference was found between the two groups when using IVUS in lumen or scaffold measurements.

A third generation magnesium-based BRS (DREAMS-3G) has been developed with larger size range, thinner struts (99/117/147 µm vs. 150 µm), and increased radial strength^24^ compared to the MgBRS used in our study. An absolute reduction in MLA was -2.4 mm^2^ (from 7.2 mm^2^ to 4.8 mm^2^ at 6-month follow-up) for the DREAMS-3G, which was comparable to our results in the standard non-compliant balloon group with an absolute reduction of -2.3 mm^2^. The scoring balloon group in our study had less absolute reduction of -1.7 mm^2^. Even though, we found a significant difference in MLA between the two groups, we still revealed lumen reduction in both groups from baseline to 6-month follow-up. Lumen reduction of 25% was considerably larger than the expected 11% lumen reduction anticipated in our power calculation.

The HONEST trial^25^ comparing OCT-and angio-guided implantation with the MgBRS in a population with acute coronary syndrome found a significant reduction in MLA observed after 6 month in both groups with a relative difference of 33.2 % and 22.8 % in MLA, respectively. The mechanism behind lumen reduction may be due to additional post-dilatation in an attempt to optimize the apposition, resulting in fracture or dismantling of the scaffold hence reducing the radial strength^26^. Other mechanisms contributing to premature lumen loss after implantation of the MgBRS could be scaffold recoil, neointimal hyperplasia and impact of underlying plaque morphology and vessel remodeling^5^. The pattern of remodeling, with significant correlation between change in lumen area and change in vessel area, but not between change in lumen area and plaque area, indicated vessel reduction and not plaque increase as the overall reason for lumen reduction. The pattern of remodeling was similar in the two groups, but the overall magnitude of vessel reduction causing lumen reduction was larger in the standard non-compliant balloon group compared the scoring balloon group. Our results reported significantly more decrease in vessel area in lesions prepared with a standard non-compliant balloon, which was not seen in the lesions pre-dilated with the scoring balloon. This indicates that negative remodeling and vessel shrinkage may be contributing factors for lumen loss in our study in the standard non-compliant balloon group. In the ABSORB Cohort B trial, dynamics of the vessel wall was investigated with IVUS after implantation the everolimus-eluting bioresorbable ABSORB scaffold. They reported no evidence of late recoil, but enlargement of the vessel, lumen and scaffold area up to three years after implantation^27^. The early resorption of the MgBRS with fast loss of radial force has been suggested as a limiting factor to the device, and must be investigated further^5^. The extent of scaffold recoil is a balance between elastic recoil and radial strength and can be affected by the fibrotic plaque in the coronary artery in the treated segment^5^. Optimal pre-dilatation with a more aggressive lesion preparation could result in a better vascular healing and less lumen reduction^11^. More lipid-rich plaques have been associated with less lumen loss after implantation of the MgBRS, whereas the constrictive vascular forces and rigidity of fibrotic plaque may facilitate lumen reduction^5^. Patients with acute coronary syndrome tend to have lesions with more lipid-rich plaque and positive remodeling compared to our population of patients with stabile coronary syndrome, which could explain more lumen reduction than expected in the current study.

Percentage of post-procedure malapposed struts was small in our study in both groups (1.46% for scoring balloon group and 4.57% for the standard non-compliant balloon group). As shown in previous trials^5, 19, 20, 22^, most struts will not be visible after 6 months, due to the fast scaffold absorption. Even though we found up to 43% of the scaffolds with malapposition had follow-up, the percentage of malapposed struts and malapposition volume was low. Significantly less malapposition was present in the scoring balloon group compared to the standard non-compliant balloon group, which contributes to the assumption of better vascular healing after lesion preparation with a scoring balloon. To determine if these findings are a part of the natural healing process needs longer follow-up time.

Despite reported lumen loss after implantation of the MgBRS in various intravascular imaging studies^19, 22^, the clinical performance is still deemed safe and efficient in several studies. Registries have reported safety and efficacy with low 1-year TLF rates of 3.3-5.4% and stent thrombosis rates of 0.5%, and TLF of 7.8% and scaffold thrombosis of 0.5% up to 24 months after implantation^9, 28,29^. A registry study found no difference in 24-month clinical outcomes between patients with acute vs. stable coronary syndromes who were treated with a MgBRS^30^. Only few studies have compared the MgBRS to DES, for example the MAGSTEMI trial (MAGnesium-based bioresorbable scaffold in ST-segment Elevation Myocardial Infarction) that showed a significantly higher TLF rate in the MgBRS group after 1 year in a ST-segment elevation myocardial infarction population^10^. However, a retrospective cohort reported similar 1-year clinical outcome comparing the MgBRS to a biodegradable polymer DES in a non-ST-segment elevation myocardial infarction cohort ^31^. More randomized controlled trials with long-term follow-up are needed to fully illuminate the clinical benefits or disadvantages between the new generation BRS and traditional DES.

## Limitations

There are some potential limitations to this study. The study was not powered to correlate clinical endpoints with OCT and IVUS findings. The study was conducted during the COVID-19 pandemic and was furthermore challenged by nurse strike and delivery problems of OCT catheters, why the inclusion period was unexpectedly prolonged. Also, the patient and lesion selections were influenced by limited available scaffold sizes.

## Conclusion

After 6 months, lesion preparation with a scoring balloon, compared to a standard non-compliant balloon, prior to implantation of a MgBRS resulted in larger MLA, no remodeling and less malapposition, whereas negative remodeling was seen in the standard non-compliant balloon group.

## Data Availability

The data that support the findings of this study are available on request from the corresponding author (KNH)

## Non-standard Abbreviations and Acronyms

BRS: Bioresorbable scaffold
DAPT: Dual antiplatelet therapy
DES: Drug-eluting stents
EEM: External elastic membrane
IVUS: Intravascular ultrasound
MLA: Minimal lumen area
MgBRS: Magnesium-based Magmaris bioresorbable scaffold
NOAC: Novel oral anticoagulant
OCT: Optical coherence tomography
OPTIMIS: Optimal Pre-dilatation Treatment before Implantation of a Magmaris bioresorbable scaffold In coronary artery Stenosis
PCI: Percutaneous coronary intervention

